# Rationale and Design of the BECA Project: Smartwatch-based Activation of the Chain of Survival for Out-of-Hospital Cardiac Arrest

**DOI:** 10.1101/2023.11.24.23298765

**Authors:** Roelof G. Hup, Emma C. Linssen, Marijn Eversdijk, Bente Verbruggen, Marieke A.R. Bak, Mirela Habibovic, Willem J. Kop, Dick L. Willems, Lukas R.C. Dekker, Reinder Haakma, Carlijn A. Vernooij, Tom A. Kooy, Hanno L. Tan, Rik Vullings

## Abstract

**Introduction:** Out-of-hospital cardiac arrest (OHCA) is a major health problem, and the overall survival rate is low (4.6%-16.4%). The initiation of the current chain of survival depends on the presence of a witness of the OHCA, which is not present in 29.7%-63.4% of the cases. Furthermore, a delay in starting this chain is common in witnessed OHCA. This project aims to reduce morbidity and mortality due to OHCA by developing a smartwatch-based solution to expedite the chain of survival in the case of (un)witnessed OHCA.

**Methods and analysis:** Within the BECA (BEating Cardiac Arrest) project, we aim to develop a demonstrator product that can detect OHCA using photoplethysmography and accelerometer analysis, and autonomously alert emergency medical services. A target group study will be performed to determine who benefits the most from this product. Furthermore, several clinical studies will be conducted to capture or simulate data on OHCA cases, as to develop detection algorithms and validate their diagnostic performance. Volunteers will be asked to simulate OHCA by interrupting radial arterial and venous blood flow by blood pressure cuff inflation while lying still. Data will also be captured during cardiac electrophysiologic and implantable cardioverter defibrillator (ICD) testing procedures. In addition, patients at risk for OHCA are recruited to acquire measurements over a longer period. Moreover, studies on psychosocial and ethical acceptability will be conducted, consisting of surveys, focus groups, and interviews. These studies will focus on end-user preferences and needs, to ensure that important individual and societal values are respected in the design process.

**Ethics and dissemination:** Ethical approval or waivers will be sought from the research ethics committees of the different institutions. Written informed consent will be obtained from the participants of all studies. Study findings will be submitted to international peer-reviewed journals and will be presented at international scientific conferences.

**STRENGTHS AND LIMITATIONS:** 

**Strengths:**

- This research project aims to develop unobtrusive technology that could save lives by autonomously alerting emergency medical services in case of out-of-hospital cardiac arrest.
- The project includes a broad range of aspects to maximize the technology’s chance of adoption: clinical, technical, psychological, and ethical.
- The project aims for the inclusion of a wide and diverse research sample and the involvement of different stakeholder groups to minimize bias and ensure accessibility for everyone in society.

**Limitations:**

- Smartwatch-obtained data of OHCA cases is scarce and hard to acquire: the more realistic the data is, the more difficult its acquisition is.

## INTRODUCTION

Sudden cardiac arrest (SCA) is a major health problem and occurs in individuals of all ages, sexes, ethnicities, and socioeconomic positions. Most SCA events occur outside hospitals (out-of-hospital cardiac arrest; OHCA). Across several national and regional registers, OHCA incidence in 2017 is estimated at 40.8 to 100.2 individuals per 100,000 population, with a survival rate of 4.6% to 16.4% [1]. In the Netherlands, a survival rate of 18% has been reported [2].

The current chain of survival starts with early recognition and alarming, after which basic and advanced life support is provided by first responders (police, firefighters, citizen responders) and emergency medical services (EMS) respectively, depending on the country or region [3]. It is essential that cardiopulmonary resuscitation and defibrillation are started as early as possible to increase the chance of survival with good neurological outcome [4, 5].

Currently, several challenges are present in this chain of survival. The initiation of the chain of survival depends on a witness of the OHCA, as victims themselves become unconscious within seconds after circulatory arrest and cannot ask for help. Without a witness, the chances of survival are practically zero [6]. Globally, in 29.7% to 63.4% of the OHCA cases, no witnesses are present [1]. In the Netherlands, approximately 40% of the cases go unwitnessed [7]. Even in witnessed OHCA, a delay in calling the emergency services is common and is associated with an increased mortality risk [8]. Furthermore, precise knowledge about the time of collapse is generally not available, although the duration of a cardiac arrest is an important predictor of the outcome of the resuscitation and this information could help caregivers in deciding on their treatment strategy [8].

In the early 2000s, it was already reasoned that a monitoring device could solve these challenges [9]. Several attempts have been made since then. In 2011, the *Wriskwatch* was introduced to detect pulselessness using a mechanical plethysmograph [10]. In the same year, a remote monitoring system for lethal arrhythmias was developed, comprising a sensor patch capturing ECG, body surface temperature, and acceleration [11]. In 2018, the *Heart Sentinel App* was tested, which uses input data from commercial ECG straps to detect heart rate irregularities [12]. Furthermore, the *iBeat Heart Watch* was introduced as a commercial blood flow monitoring smartwatch [13]. A recent study has also used microphones from smartphones and smart speakers to detect agonal breathing, a common symptom of cardiac arrest [14].

In a recent systematic review of OHCA detection technologies, several shortcomings have been mentioned regarding these earlier attempts [15]:

- Testing conditions were idealized by using high-quality data, not accounting for measurement variability, suboptimal device contact, or data loss.
- Sensitivity was generally tested on simulated data or annotated databases, while specificity was tested through direct sensor deployment, creating a mismatch in measurement environment characteristics.
- A focus was placed on the optimization of sensitivity generally lowers the diagnostic specificity, which may lead to many false alarms overburdening EMS operations.
- Integration in the current EMS chain of care was not investigated carefully.
- No follow-up studies have been conducted, which may be due to a lack of user interest, a lack of reliable methods to validate OHCA detection, or due to market conditions.

In the BECA (BEating Cardiac Arrest) project, we aim to build upon previous work by developing a demonstrator of a concept based on a smartwatch, which can be used for non-invasive and long-term continuous monitoring to detect SCA and to autonomously alert EMS, first responders, and bystanders in case of OHCA. The demonstrator will use photoplethysmography (PPG) and accelerometry to detect SCA. We intend to maximize diagnostic performance and generalization by developing new detection algorithms based on newly acquired clinical data. In contrast to previous works, our new data will be of added value by acquiring the data in a more day-to-day setting, instead of controlled (hospital) environments. We aim to establish an acceptable balance between sensitivity and specificity by interviewing various stakeholders, such as physicians, first responders, and patients at increased risk of SCA. Furthermore, we will investigate how to integrate this detection technology into the Dutch EMS system. Lastly, broader end-user preferences will also be considered in the design process by focusing on end-user needs and ethical, psychosocial, and legal aspects, such that the acceptance, compliance, and value-based implementation options of this newly developed product are safeguarded.

By investigating the technical, clinical, psychological, and ethical aspects of using our innovative technology, this project aims to increase the chance of adoption by a broad range of users in society to ultimately raise OHCA survival rates. To accomplish this goal, the BECA project will be conducted by a multi-disciplinary consortium consisting of academic partners from the areas of cardiology, engineering, psychology, and ethics, as well as a large MedTech company and a technology provider for citizen responder systems.

## METHODS AND ANALYSIS

In this project, we aim to comprehensively develop a demonstrator, showcasing a possible concept of a minimum viable product. Here, the earlier mentioned multi-disciplinary aspects are included in the research scope. In this section, we discuss the multiple studies that will be conducted on the validation and optimization of diagnostic accuracy and on maximizing the psychosocial and ethical acceptability of our demonstrator, as well as the successful integration of the demonstrator into the chain of survival.

### Study on the initial target group

This analysis aims to identify the initial target group for BECA, i.e., the persons who are likely to benefit the most from the smartwatch. We define these persons as patients who suffered OHCA in the absence of a witness. Identification of the initial target group may aid in designing the smartwatch. To identify the characteristics of these patients, we will use data from the ARREST database [16] which was built by consortium partner Amsterdam UMC. The ARREST database contains information on all EMS-attended OHCA cases that occur in the province of North Holland in the Netherlands. The data collection started in 2005 and is ongoing, which led to more than 40.000 included cases until now. The database contains information on the circumstances of the OHCA (e.g., presence of a witness, time logs of EMS alerting, dispatch, and arrival) as well as characteristics of the patient (e.g., demographics, comorbidities). Next to this, we will use data from Statistics Netherlands (e.g., ethnicity, income, socioeconomic position). With this information, we will characterize the group of OHCA patients who suffered from an unwitnessed cardiac arrest, both EMS-attended and not EMS-attended. By characterizing these groups, we can determine what our initial target group will look like, for example in terms of sex, age, and socioeconomic position.

### The smartwatch and its integration into the chain of survival

In this project, we aim to develop a concept demonstrator based on a clinical data logger that captures PPG and accelerometric signals at the wrist. ECG sensors were considered, but are not practical for long-term continuous monitoring. ECG patches may cause skin irritation and need regular replacement [17], while ECG-equipped wristwatches require the wearer to place the finger of their opposite hand on the smartwatch, which is not feasible for long-term monitoring and impossible for unconscious OHCA victims. As PPG does not suffer from these shortcomings, we chose to study a PPG-based system over an ECG-based system.

As part of the concept demonstrator, the clinical data logger will be equipped with user controls and will allow for connectivity to the emergency response infrastructure. Whenever the demonstrator is worn and a circulatory arrest is detected, it should raise an alarm to allow the wearer to cancel the alarm, as to minimize the number of false alerts in the EMS infrastructure. If this alarm is not canceled within a certain time frame, the EMS are warned. This process is visualized in Figure 1.

**Figure 1.**
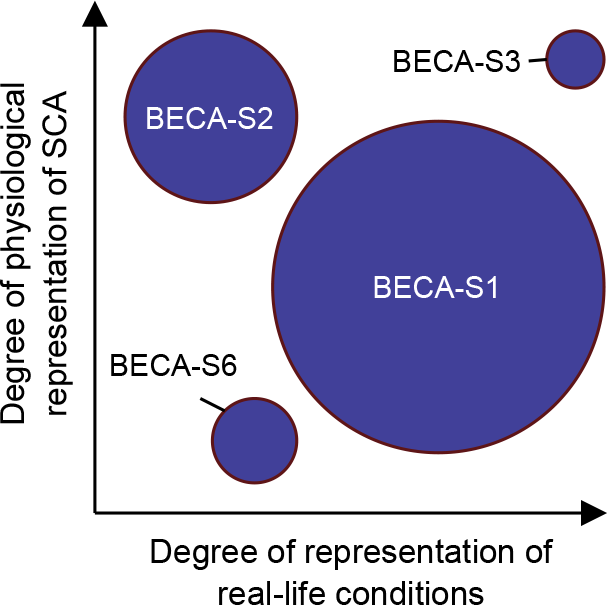
Decision flowchart for the final product. Several steps are taken before a remote alarm is triggered.

If the demonstrator shows success, a minimum viable product can be developed based on the findings of the BECA project. For such a product, we envision a solution where the concept demonstrator is extended with mobile connectivity, GPS localization functionality, and full integration into the already existing Dutch citizen responder system *HartslagNu*.

### Studies on validation and optimization of diagnostic accuracy

To develop the demonstrator and evaluate its performance, we will conduct several studies with several groups of patients and volunteers. Four studies will be conducted to develop and validate an algorithm for smartwatch based OHCA detection, and one study will focus on the actual number of false alarms that will reach the EMS infrastructure when users get the chance to cancel alarms.

As capturing sufficient real OHCA data would require unreasonably large studies for our current stage and purpose, most of the studies aim to approximate some of the characteristics of OHCA, i.e., the physiological nature of SCA and the difference in body movements between normal situations and OHCA cases. The trade-off between the OHCA occurrence and the degree of representativeness based on the earlier-mentioned characteristics is shown in Figure 2, which shows that studies with higher occurrence tend to be less representative of either one of the OHCA characteristics. We presume that the combination of these studies represents real OHCA cases sufficiently well and provides enough data to develop and validate a demonstrator.

**Figure 2.**
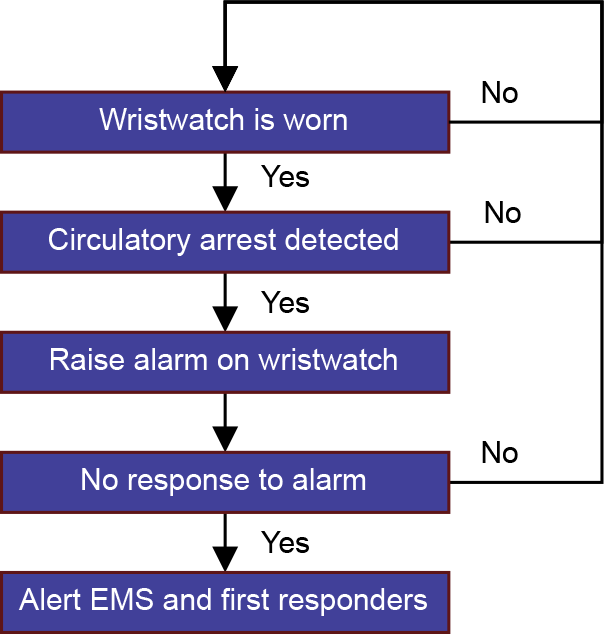
Representativeness of the different studies with regard to SCA physiology and real-life conditions. Circle size indicates the relative amount of (simulated) OHCA cases for that study.

#### BECA-S1: Repeated simulation of OHCA in healthy volunteers

In this study, 60 healthy volunteers will be selected from the general population. The volunteers are asked to wear a clinical data logger continuously for 28 days. The data logger will capture continuous PPG and accelerometric measurements at the wrist. During the study period, the volunteers will be asked to simulate 25 cardiac arrest episodes by laying down and staying as still as possible, while inflating a blood pressure cuff on the watch-wearing arm, which inhibits pulsatile blood flow. They will be instructed to inflate the blood pressure cuff to 200 mmHg and keep it inflated for 10 seconds. All volunteers will be given the same pressure cuffs to standardize across the cohort. The timestamps of these simulated episodes will be marked in a diary, which serves as ground truth for the development and validation of SCA detection algorithms. This method simulates real-life conditions, such as movement patterns, quite well, but it simulates the physiological characteristics of SCA less accurately, as the inflation of the cuff causes blood pooling in the arm and thus a higher PPG baseline [18].

#### BECA-S2: Induced ventricular tachycardia and fibrillation in a clinical setting

The second study will follow patients who are scheduled for procedures in which ventricular fibrillation/tachycardia (VF/VT) - the cardiac arrhythmia that underlies most cases of OHCA - is expected or induced. We will select 30 patients undergoing an electrophysiologic study with programmed electrical stimulation to induce VF/VT, e.g., for catheter ablation. Furthermore, we will select 15 patients who are scheduled for implantation of a first subcutaneous implantable cardioverter defibrillator (S-ICD) and undergo S-ICD testing. During both types of procedures, patients will wear a clinical data logger. Furthermore, ECG will be captured to identify VT/VF episodes. While this study approximates circulatory arrest, the real-life conditions of OHCA are not represented, as the patients will be sedated and lying still.

#### BECA-S3: Real SCA in patients with ICDs

The third study will be conducted with 30 patients who have an ICD because of an increased risk of VT/VF occurrence, e.g., with a history of recurring episodes of VT-/VF-induced circulatory arrest. These patients will wear the clinical data logger for 9 consecutive months and keep a diary of VT/VF episodes and their timestamps. Electrograms from the ICD will be captured to verify the nature and occurrence of the episodes. The data acquired with this study may represent real OHCA cases the best, as the only difference between OHCA victims and VT/VF episodes in this patient group is the presence of an ICD. A limitation is the unpredictability of ICD shocks in the study population, leading to uncertainty about whether this study will acquire enough data on cardiac events.

#### BECA-S6: Simulated OHCA in healthy volunteers

To quickly acquire usable data and extend the population of BECA-S1, this study will ask 100 volunteers to wear clinical data loggers on both wrists for approximately one hour while performing their daily activities. After this hour, the investigator instructs the volunteer to stay still and inflates a blood pressure cuff on one of the arms to 200 mmHg of pressure for a maximum duration of 20 seconds. Similar to BECA-S1, real-life conditions are simulated quite well, but the physiological characteristics of SCA are simulated less accurately.

#### BECA-S7: User response time and rate for false alarm suppression

As the previous studies solely focus on the performance of the detection algorithm, the ability of users to cancel false alarms creates uncertainty about how many false alarms will actually reach the EMS infrastructure. In this study, 30 volunteers of varying age groups will be asked to wear a programmable smartwatch for 10 hours. Within this time frame, 18 alarms are produced, which can be audible alarms, tactile (vibration) alarms, or a combination of both. Volunteers are instructed to turn off these alarms as quickly as possible by touching the screen of the watch. The watch itself records the time between activation and deactivation of the alarm. Together with volunteer activity logs and surveys, we will perform qualitative and quantitative analysis to determine how long the time window should be in which users of the minimum viable product are able to cancel a false alarm.

#### Existing datasets

We aim to use existing datasets containing PPG and/or accelerometer data to increase the robustness of the demonstrator to different measurement circumstances and sensors. Most publicly available datasets do not contain fully representative OHCA episodes, but will represent OHCA in either the physiology of SCA or the real-life measurement circumstances. As the number of SCA episodes in these datasets is low, our clinical trials will complement these.

#### Data analysis

The acquired data, combined with existing datasets, will be used to develop a binary classifier model that determines for a certain small window of time (e.g., 100 ms – 10 s) whether a circulatory arrest has occurred in that window. PPG and accelerometer signals within this window are the main data of interest, but they can be supplemented by prior signal data, e.g., from minutes or hours before the event. Different window sizes will be examined to determine the optimal balance between classification performance and the speed of classification.

Due to the relatively low number of patients and volunteers, we will evaluate this model using Leave-One-Patient-Out Cross-Validation and present Receiver Operator Characteristics curves and their Area Under the Curve based on the classifier threshold. We aim to present how the classifier will perform in real life, e.g., by indicating the number of false alarms per month for a given sensitivity.

### Studies to maximize psychosocial and ethical acceptability

In order to develop a psychologically and ethically acceptable product, several studies will be performed on various groups of stakeholders with respect to OHCA. We aim to deliver a user-centered template for the technology at the end of the project. These studies will focus on end-user preferences and needs and will empirically motivate the ethical and legal aspects of the technology (‘ethics by design’), such as preventing privacy breaches or the discriminatory use of data [19]. This approach will ensure that important individual and societal values are respected, and it will aim to increase satisfaction with care and decrease possible psychological distress associated with wearing the smartwatch. In these studies, we aim for the inclusion of a diverse population in terms of age, sex and gender, ethnicity, and socio-economic status, to accumulate a broad set of values and perspectives related to the use of smartwatch-based technology in (unwitnessed) OHCA. Particular attention will be paid to people with elevated distress levels (or those prone to experiencing distress or other unintended psychological consequences) as this subgroup might require a different approach to ensure the uptake of the technology.

#### BECA-S4: Cross-sectional study on end-user preferences

The first study focusing on psychosocial and ethical acceptability will be a cross-sectional study using a questionnaire to be completed by participants from several stakeholder groups (e.g., patients, their significant others, healthy volunteers, and care workers). A full overview of the targeted groups, a rationale behind their inclusion and the desired sample sizes can be found in the Supplemental Materials. Questionnaire topics will be tailored to specific group circumstances, but they will also cover the same general topics of the project. Topics include (1) demographic variables; (2) personal perspectives on heart disease and cardiac arrest; (3) questions regarding the smartwatch on user expectancy, reliability, privacy, handling incidental findings, and open-ended questions for free text input; (4) perceived stress and psychological trait characteristics; (5) technology in health care in general and (6) personal health and lifestyle factors. Questions for care providers will cover topics including (1) practical concerns after the alarm is raised (e.g., opening locked doors when no witness is present inside); (2) monitoring and handling incidental findings; (3) acceptable thresholds regarding false alarms and (4) other design requirements from the perspective of the care provider using open-ended questions. The reliability of the questionnaires will be assessed by calculating Cronbach’s alpha, while validity will be assessed by examining the internal factor structure.

#### BECA-S5: Focus groups and interviews on end-user preferences

The second study focusing on psychosocial and ethical acceptability will consist of focus groups and individual interviews to further elaborate on the findings of the cross-sectional study. Each focus group will consist of four to eight participants from the target stakeholder groups, while the interviews are held individually or together with a partner. During these sessions, the interviewer will discuss various psychological and ethical challenges related to the project, such as expected psychological distress while wearing the smartwatch, consent, autonomy, privacy, justice and accessibility, responsibility and liability, and yet unforeseen ethical and psychological issues. We will determine the number of sessions based on an iterative evaluation of the level of data saturation throughout the interviews. We will start with three to five sessions per target group, where we will write a short summary afterward in a logbook. After each set of sessions, the similarity between these summaries will be evaluated, which will then influence our decision to organize additional sessions or whether a certain level of saturation has already been reached. With this approach, we aim to gather enough data in a comprehensive and conclusive manner. In order to increase the validity of this study we will also include a member check which allows participants to analyze the summaries and comment on them.

#### Data analysis

The data from the questionnaires will be analyzed using both descriptive and inferential statistics. In general, we will assess differences in age, sex and gender, socio-economic status, (perceived) health, anxiety, stress, coping style, and other personality traits related to the smartwatch-specific questionnaire items. This will be done in an exploratory manner, to find out which specific topics are most important for different target groups when it comes to the willingness to use the technology. Findings from this analysis will be validated by including them as topics in the focus groups and interviews, to assess similarity in findings. The answers from the focus groups and interviews will be analyzed through a process of inductive coding, where labels of recurring themes will be created based on the full transcripts. By counting the occurrence of similar codes, we will identify the common themes in the data.

## Patient and public involvement

Patients and the public were not directly involved in the initial design, conduct, reporting, or dissemination of the research plans, but their input was gathered through multiple user committee meetings. During these meetings, researchers consulted representatives of patient organizations, technology developers, and governmental institutions for their input on the study protocol of the project. Additionally, stakeholders will also be involved in the studies on psychosocial and ethical acceptability, in which their perspectives are considered important study outcomes as well as recommendations for shaping subsequent research.

## Supporting information

Supplemental Material Overview of stakeholder groups

## Data Availability

There are no data produced in the present work.

## ACKNOWLEDGMENTS

We appreciate the helpful suggestions of Dr. Patrick Schober on an earlier version of this manuscript.

## ETHICS AND DISSEMINATION

The studies involve human participants and need to comply with laws and ethical regulations. Where applicable, approval or waivers will be sought from the research ethics committees of the different institutions where the research takes place. Before taking part in one of the studies, participants must have given informed consent to participate. All studies will be performed in accordance with the Declaration of Helsinki, and we will use the International Conference on Harmonization Good Clinical Practice (ICH-GCP) as guidance throughout all study-related activities. A data management plan has been developed to ensure the privacy of the study participants. The findings of the studies will be submitted to international peer-reviewed journals and will be presented at international scientific conferences.

## AUTHOR’S CONTRIBUTIONS

All authors participated in conceiving the BECA project. Emma C. Linssen, Roelof G. Hup, Marijn Eversdijk, and Bente Verbruggen drafted the manuscript. Emma C. Linssen and Roelof G. Hup contributed equally to this manuscript. All authors substantially revised and approved its contents and agreed to be accountable for the work.

