## Supplemental Material Overview of stakeholder groups for "Rationale and Design of the BECA Project: Smartwatch-based Activation of the Chain of Survival for Out-of-Hospital Cardiac Arrest"

### Supplemental Material: Overview of stakeholder groups in cross-sectional study on end-user preferences (BECA S4)

| **Stakeholder group (Sample size)** | **Rationale** |
|---|---|
| Patients with an ICD due to a history of often recurring episodes of VT-/VF-induced circulatory arrest (N=30). | These will be the same participants as in *BECA-S3: Real SCA in patients with ICDs*. Although the technology is not intended to be used by patients with an ICD, patients in this group often have a lot of experience with living with heart disease, which might give valuable insights in terms of design requirements. |
| Patients without an ICD but at high risk of experiencing OHCA, as a result of cardiac arrhythmias or coronary artery diseases (N=30). | The patient group will be studied to establish what key considerations must be made to increase the willingness to continuously, or at least as much as possible, wear the technology. Both patients with congenital heart disease (Long QT, Brugada, hypertrophic cardiomyopathy, dilated cardiomyopathy and arrhythmogenic right ventricular cardiomyopathy) as patients with an earlier myocardial infarction or dilated cardiomyopathy will be included. |
| Family members or significant others (N=30), neighbors or friends (N=30) of patients at high risk of cardiac arrest. | Our technology could not only impact the lives of the patients themselves, but also the lives of their significant others. They might for example be more comfortable with the idea to leave the patient unwitnessed for some time, but might also have privacy concerns related to sharing their home address. In addition, they might also play an important role in finding the specific location of the patients or opening the door for medical personnel. We will study these and other relevant considerations related to the perspective of the patient's significant others. |
| Healthy volunteers, selected from a group of citizen responders in the *HartslagNu* program (N=60). | These will be the same participants as in *BECA-S1: Repeated simulation of OHCA in healthy volunteers*. This group is selected because of their experience with acting as first responders. |
| Healthy volunteers, selected from the general population. We especially aim for an equal distribution in terms of sex and gender, in combination with ethnic and socio-economic diversity (N=800). | Fundamental ethical questions will be studied, such as the proportionality to which this healthy population would benefit from the technology versus the risks, but also psychological questions, such as what characterizes individuals who are willing to wear the technology and what are the psychological barriers to wearing the technology. The high sample size can be attained by performing the study as part of a yearly research project at Tilburg University, with sample sizes ranging between 1000 and 2000 participants. |
| Care workers: Emergency medical service responders (N=20), dispatch center employees (N=20), basic and advanced life support trainers (N=20). | These care workers all play essential roles in the execution of successful life support, making their opinions important for successful implementation of the BECA device into the chain of survival. Questions will also focus on acceptable thresholds with regard to the trade-off between false positives (alarm raised while cardiac arrest is absent) and false negatives (no alarm raised during cardiac arrest). |
| Physicians and other medical personnel, including cardiologists (N=20), neurologists, anesthesiologists and intensivists (N=20), nurses, general physicians and rehabilitation specialists (N=20). | Questions will focus on topics such as which specific patient groups should use the technology and how incidental findings should be handled. |
